# Aberrant perception of environmental volatility during social learning in emerging psychosis

**DOI:** 10.1101/2023.02.02.23285371

**Authors:** Daniel J. Hauke, Michelle Wobmann, Christina Andreou, Amatya Mackintosh, Renate de Bock, Povilas Karvelis, Rick A. Adams, Philipp Sterzer, Stefan Borgwardt, Volker Roth, Andreea O. Diaconescu

**Author notes:** Corresponding author: Daniel J. Hauke, 90 High Holborn, 1^*st*^ Floor, London, WC1V 6LJ, United Kingdom.

## Abstract

Paranoid delusions or unfounded beliefs that others intend to deliberately cause harm are a frequent and burdensome symptom in early psychosis, but their emergence and consolidation still remains opaque. Recent theories suggest that aberrant prediction errors lead to a brittle model of the world providing a breeding ground for delusions. Here, we employ a Bayesian approach to test for a more unstable model of the world and investigate the computational mechanisms underlying emerging paranoia.

We modelled behaviour of 18 first-episode psychosis patients (FEP), 19 individuals at clinical high-risk for psychosis (CHR-P), and 19 healthy controls (HC) during an advice-taking task, designed to probe learning about others’ changing intentions. We formulated competing hypotheses comparing the standard Hierarchical Gaussian Filter (HGF), a Bayesian belief updating scheme, with a mean-reverting HGF to model an altered perception of volatility.

There was a significant group-by-volatility interaction on advice-taking suggesting that CHR-P and FEP displayed reduced adaptability to environmental volatility. Model comparison favored the standard HGF in HC, but the mean-reverting HGF in CHR-P and FEP in line with perceiving increased volatility, although model attributions in CHR-P were heterogeneous. We observed correlations between shifts in perceived volatility and positive symptoms generally as well as with frequency of paranoid delusions specifically.

Our results suggest that FEP are characterised by a different computational mechanism – perceiving the environment as increasingly volatile – in line with Bayesian accounts of psychosis. This approach may prove useful to investigate heterogeneity in CHR-P and identify vulnerability for transition to psychosis.

## 1 Introduction

Paranoid delusions are commonly defined as unfounded beliefs that others intend to deliberately cause harm (Freeman and Garety, 2000) and they are a frequent symptom in early psychosis occurring in about 50-70% of first-episode-psychosis patients (FEP) (Freeman, 2007; Freeman and Garety, 2014; Sartorius et al., 1986). While paranoid delusions are a key symptom of schizophrenia, they are also present in the general population (Freeman et al., 2005; Wellstein et al., 2020) and are frequently reported in other psychotic disorders and affective disorders, such as bipolar disorder and depression (Appelbaum et al., 1999). Importantly, paranoid delusions are a heavy burden for those afflicted by them as they are associated with more frequent suicidal ideation in the general population (Freeman et al., 2011) and higher suicide risk in patients (Fenton et al., 1997; Saarinen et al., 1999). Despite an urgent clinical need to address these symptoms, the emergence and consolidation of paranoid delusions remain a subject of debate. Recent cognitive theories suggest that aberrant salience caused by overly precise prediction errors (PEs) – possibly mediated through dopaminergic signaling – lead to a brittle model of the world providing a breeding ground for delusions to form (Kapur, 2003; Howes and Kapur, 2009; Corlett et al., 2010; Winton-Brown et al., 2014; Diaconescu et al., 2019). It has been proposed that these aberrantly salient PEs could then be explained away by adopting more abstract higher order beliefs that may take the form of delusions (Kapur, 2003; Corlett et al., 2010; Sterzer et al., 2018).

Here, we pursue a Bayesian approach that enables us to formalize the concept of aberrant salience. We will first discuss aberrant salience in a non-hierarchical framework and then proceed to a hierarchical framework using a hierarchical Bayesian model of learning (Mathys et al., 2011, 2014) to derive competing computational mechanisms that are tested in this study. When adopting a Bayesian framework, aberrant salience can be understood as reduced uncertainty (i.e., variance) or increased precision (inverse of uncertainty) that up-weighs incoming sensory information (Stephan et al., 2006; Fletcher and Frith, 2009; Corlett et al., 2009, 2010; Adams et al., 2013; Diaconescu et al., 2019). In a non-hierarchical model, aberrant salience would be expressed in relatively increased precision associated with the likelihood or reduced precision associated with the prior distribution (e.g., see Sterzer et al. (2018)).

However, for example Fletcher and Frith (2009) have argued that beliefs may better be conceptualised in a hierarchical manner. Assuming a hierarchical structure of beliefs where the lower level corresponds to beliefs about sensory information and the higher level to beliefs about the volatility of the environment and further assuming that beliefs can be expressed as Gaussian distributions, aberrant salience can be viewed as a ratio of precisions associated with beliefs about sensory inputs and high-level beliefs (Mathys et al., 2011, 2014; Diaconescu et al., 2019). An increase in this precision ratio will result in exaggerated belief updates or aberrantly salient PEs. From here on out we will refer to beliefs about volatility when we speak about high-level beliefs.

In line with this literature, we have recently derived different hypotheses about the emergence of delusions based on simulations (Diaconescu et al., 2019) using the Hierarchical Gaussian Filter (HGF; (Mathys et al., 2011, 2014)). Specifically, we hypothesised that prodromal stages of psychosis may be characterized by either (1) increased precision associated with incoming sensory prediction errors (2) reduced precision of high-level beliefs about the volatility of the environment or (3) a combination of the two. Furthermore, we speculated that delusional conviction during later stages of psychosis may be accompanied by a compensatory increase of precision associated with high-level beliefs about volatility that functions to explain away aberrantly salient prediction errors. Here, we test these hypotheses and investigate the computational mechanisms of emerging paranoia in early psychosis.

## 2 Methods

### 2.1 Participants

The sample comprised 19 individuals at clinical high risk for psychosis (CHR-P), 19 healthy controls (HC) that were group-matched to CHR-P with respect to age, gender, handedness, and cannabis consumption, and 18 short term medicated FEP (5.44 ± 2.79 days, median: 6, range: [0, 10]) resulting in a total sample of *N* = 56 participants. FEP were recruited from both inpatient care and the outpatient departments of the University Psychiatric Hospital (UPK) Basel, CHR-P were recruited from the Basel Early Treatment Service (BEATS) and HC via online advertisements and advertisements in public places (supermarkets, dentist clinics). All participants provided written informed consent. The study was approved by the local ethics committee (Ethikkommission Nordwest- und Zentralschweiz, no. 2017-01149) and conducted in accordance with the latest version of the Declaration of Helsinki.

### 2.2 In- and exclusion criteria

All participants were required to be at least 15 years old. Specific inclusion criteria for FEP were the diagnosis of a first psychotic episode of a schizophrenia spectrum disorder, which was assessed by the treating clinicians, and a treatment recommendation to begin antipsychotic medication issued independently of the study.

We included CHR-P who fulfilled either ultra-high risk for psychosis criteria, i.e. one or more of the following (1) attenuated psychotic symptoms (APS), (2) brief and limited intermittent psychotic symptoms (BLIP), (3) a trait vulnerability in addition to a marked decline in psychosocial functioning also referred to as genetic risk and deterioration syndrome (GRD), assessed with the Structured Interview for Prodromal Symptoms (SIPS; Miller et al. (2003)); or basic symptom criteria, (Klosterkötter et al., 2001; Schultze-Lutter, 2009) i.e., cognitive-perceptive basic symptoms (COPER) or cognitive disturbances (COGIDS) (assessed with the Schizophrenia Proneness Instrument, adult version (SPI-A; Schultze-Lutter et al. (2007)) or the Schizophrenia Proneness Instrument, child and youth version (SPI-CY; Schultze-Lutter and Koch (2010)), assessed by experienced clinical raters.

Exclusion criteria for all three groups were previous psychotic episodes, psychotic symptomatology secondary to an organic disorder, any neurological disorder (past or present), premorbid IQ *<* 70 (assessed with the Mehrfachwahl-Wortschatz-Test, Version A; Lehrl et al. (1995)), colour blindness, substance use disorders according to ICD-10 criteria (except cannabis), alcohol or cannabis consumption within 24 hours prior to measurements, and regular drug consumption (except alcohol, nicotine, and cannabis), which was assessed during the admission interview and confirmed with a drug screening before the initial measurement (assessments were postponed following a positive test until a negative test result was obtained).

FEPs whose psychotic symptoms were associated with an affective psychosis or a borderline personality disorder at the time of the measurement were excluded. Since data was collected as part of a larger study that included neuroimaging assessments, additional exclusion criteria for CHR-P and HC were contraindications for fMRI and contraindications for EEG measurements for all three groups. However, we only present behavioural results here.

### 2.3 Clinical assessment

Demographic and clinical information were assessed during an interview conducted within five days of the social learning task. This interview comprised assessment of clinical symptoms using the Positive and Negative Syndrome Scale (PANSS; Kay et al. (1987)) administered through trained clinical raters and self-assessment of paranoid thoughts (frequency, conviction and distress) using the Paranoia Checklist (PCL) (Freeman et al., 2005).

### 2.4 Task

All participants were asked to perform a deception-free and ecologically valid social learning task (Figure 1A) (Diaconescu et al., 2014, 2017), which required them to learn about the intentions of an adviser that changed over time. The task comprised two phases. In the first phase participants received *stable* helpful advice, whereas advisers intentions were changing more rapidly during a second phase, the *volatile* phase (see volatility schedule in Figure 1B). Participants were asked to predict the outcome of a binary lottery on each trial. To this end, they received information from two sources, a non-social cue displaying the true winning probabilities of the lottery, and a recommendation of an adviser (social cue) presented in form of prerecorded videos that were extracted from trials in which a human adviser either tried to help or deceive a player in a previous human-human interaction (see Diaconescu et al. (2014, 2017) for more details).

**Figure 1:**
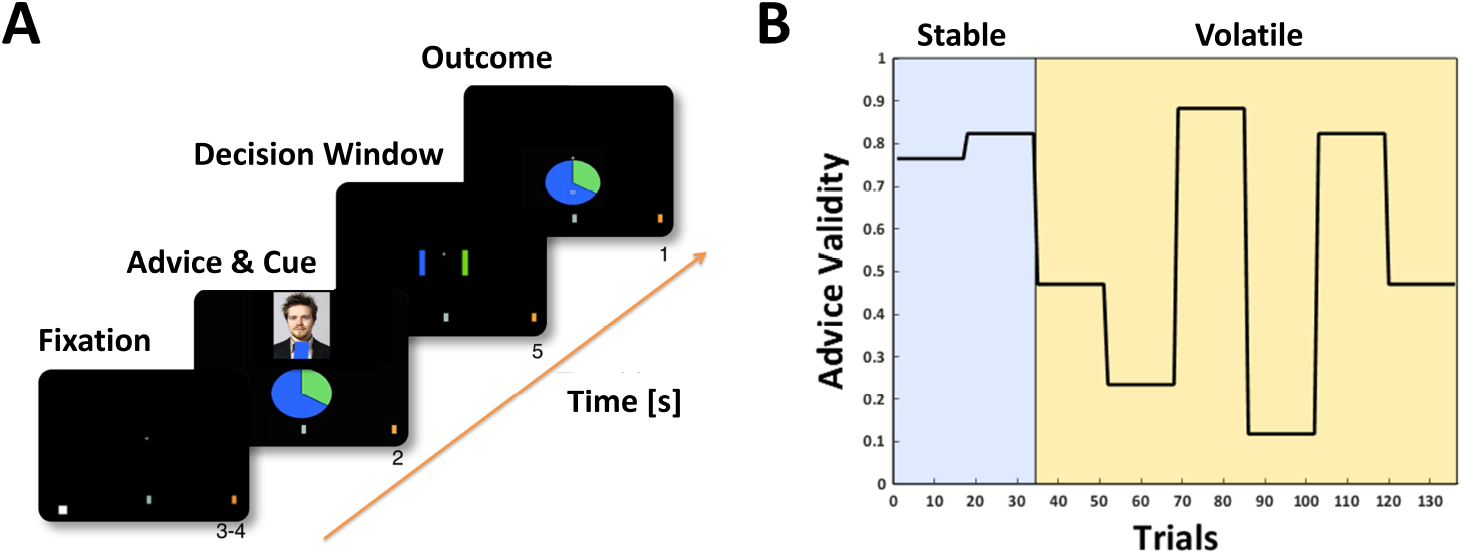
Social learning task and volatility schedule. **A** Social learning task. **B** Volatility schedule.

Participants were truthfully informed that the adviser received privileged – but not complete – information about the upcoming outcome and that inaccurate advice could be due to mistakes or that the adviser could pursuit a different agenda than the player and that the adviser’s intentions could change during the course of the experiment. We expected patients to be more sensitive to the increasing volatility of the task compared to HC.

### 2.5 Computational modelling

#### 2.5.1 Hierarchical Gaussian Filter

We modelled participants’ behaviour during the social learning task with a 3-level HGF (Mathys et al., 2011, 2014). The model comprises a perceptual model and a response model, which will be detailed below.

##### Perceptual model

The standard 3-level HGF assumes that participants infer on a hierarchy of hidden states in the world *x*_1_, *x*_2_, and *x*_3_ that cause the sensory inputs that participants perceive (Mathys et al., 2011, 2014). Participants’ inference on the true hidden states of the world 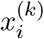 at level *i* of the hierarchy on trial *k* are denoted 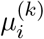. In the context of this task, the states that participants need to infer on based on the experimental inputs on each trial (non-social cue and advice) are structured as follows: The lowest level state corresponds to the *advice accuracy*. On each trial *k* an advice can either be accurate 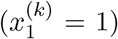 or inaccurate 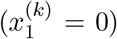. This state can be described by a Bernoulli distribution that is linked to the state at the second level 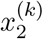 through the unit sigmoid transformation:

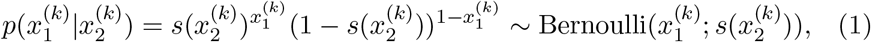

with

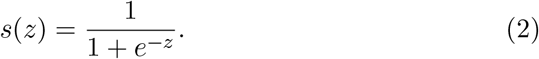

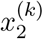 represents the unbounded tendency towards helpful advice (−∞, +∞) or the *adviser’s fidelity* and is specified by a normal distribution:

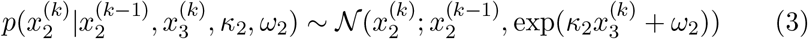

The state at the third level 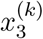 expresses the (log) volatility of the adviser’s intentions over time and is also specified by a normal distribution:

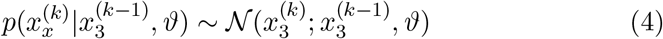

The dynamics of these states are governed by a number of subject-specific parameters, i.e., the *evolution rate* at the second level *ω*_2_, the *coupling strength* between the second and third level *κ*_2_, which determines the impact of the volatility of the adviser’s intentions on the belief update at the level below, and the evolution rate at the third level or the *meta-volatility ϑ*, which we fixed to a value of 0.5 to reduce the number of free parameters. Additional subject-specific, free parameters were the *prior expectations* before seeing any input about the adviser’s fidelity 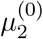 and the volatility of the adviser’s intentions 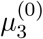 (see Table 1 for priors on all free parameters). These parameters can be understood as an individual’s approximation to Bayesian inference and provide a concise summary of a participant’s learning profile. Using a variational approximation, efficient one step update equations can be derived (see Mathys et al. (2011, 2014) for more details), which take the following form:

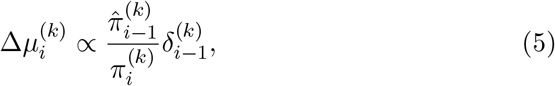

where 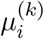 is the expectation or belief at trial *k* and level *i* of the hierarchy, 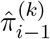 is the precision (inverse of the variance) from the level below (the hat symbol denotes that this precision has not been updated yet and is associated with the prediction before observing a new input), 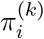 is the updated precision at the current level, and 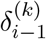 is a PE expressing the discrepancy between the expected and the observed outcome.

**Table 1:** Priors on free model parameters. Prior means and their respective variances are denoted in brackets, followed by upper bounds for parameters that were estimated in logit space: (Mean, Variance), upper bound.

|  | Equilibrium point | Coupling strength | Evolution rate | Prior expectations |  | Advice weight | Decision noise |
| --- | --- | --- | --- | --- | --- | --- | --- |
| Hypothesis I | | $\kappa_2(\text{logit}(0.5), 1), 1$ | $\omega_2(-2, 4)$ | $\mu_2^{(0)}(0, 1)$ | $\mu_3^{(0)}(1, 1)$ | $\zeta(\text{logit}(0.5), 1), 1$ | $\nu(\text{log}(48), 1)$ |
| Hypothesis II | $m_3(1, 1)$ | $\kappa_2(\text{logit}(0.5), 1), 1$ | $\omega_2(-2, 4)$ | $\mu_2^{(0)}(0, 1)$ | $\mu_3^{(0)}(1, 1)$ | $\zeta(\text{logit}(0.5), 1), 1$ | $\nu(\text{log}(48), 1)$ |

We also employed a second, modified version of the HGF (Cole et al., 2020) that assumed that learning about an adviser’s intentions was not only driven by hierarchical PE updates, but also included a mean-reverting process at the third level formalising the idea that an altered perception of volatility may underlie learning about others’ intentions. In this mean-reverting HGF, the third level can again be described by a normal distribution:

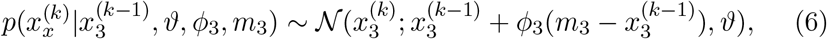

where *ϕ*_3_ represents a drift rate and *m*_3_ the equilibrium point towards which the state moves over time.

In this model, we fixed the drift rate *ϕ*_3_ to a value of 0.1 and estimated the equilibrium point *m*_3_ as a subject-specific, free parameter. Note, that changing *m*_3_ to values that are lower than the prior about the volatility of the adviser’s intentions 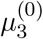 translates into reduced belief updates at all three levels of the hierarchy corresponding to perceiving the environment as increasingly stable over time (Figure 2). Conversely, if 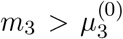, the magnitude of belief updates increases in line with a perception that the environment is increasingly volatile over time and beliefs should thus be adjusted more rapidly. Lastly, if 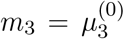, agents would revert back to their prior beliefs about environmental volatility over time (i.e., “forget” about the observed inputs). For this reason, we refer to the model as *mean-reverting* HGF analogous to an Ornstein-Uhlenbeck process in discrete time (Uhlenbeck and Ornstein, 1930). Note, that introducing this drift allows to model an altered perception of volatility that manifest not only during the first trials as changes in prior uncertainty 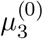 would induce (see simulations in the Supplement), but rather enables a more nuanced characterization of changes that occur *within* the experimental session. Its effect also impacts belief formation at lower levels and simulated responses more strongly (see Supplement).

**Figure 2:**
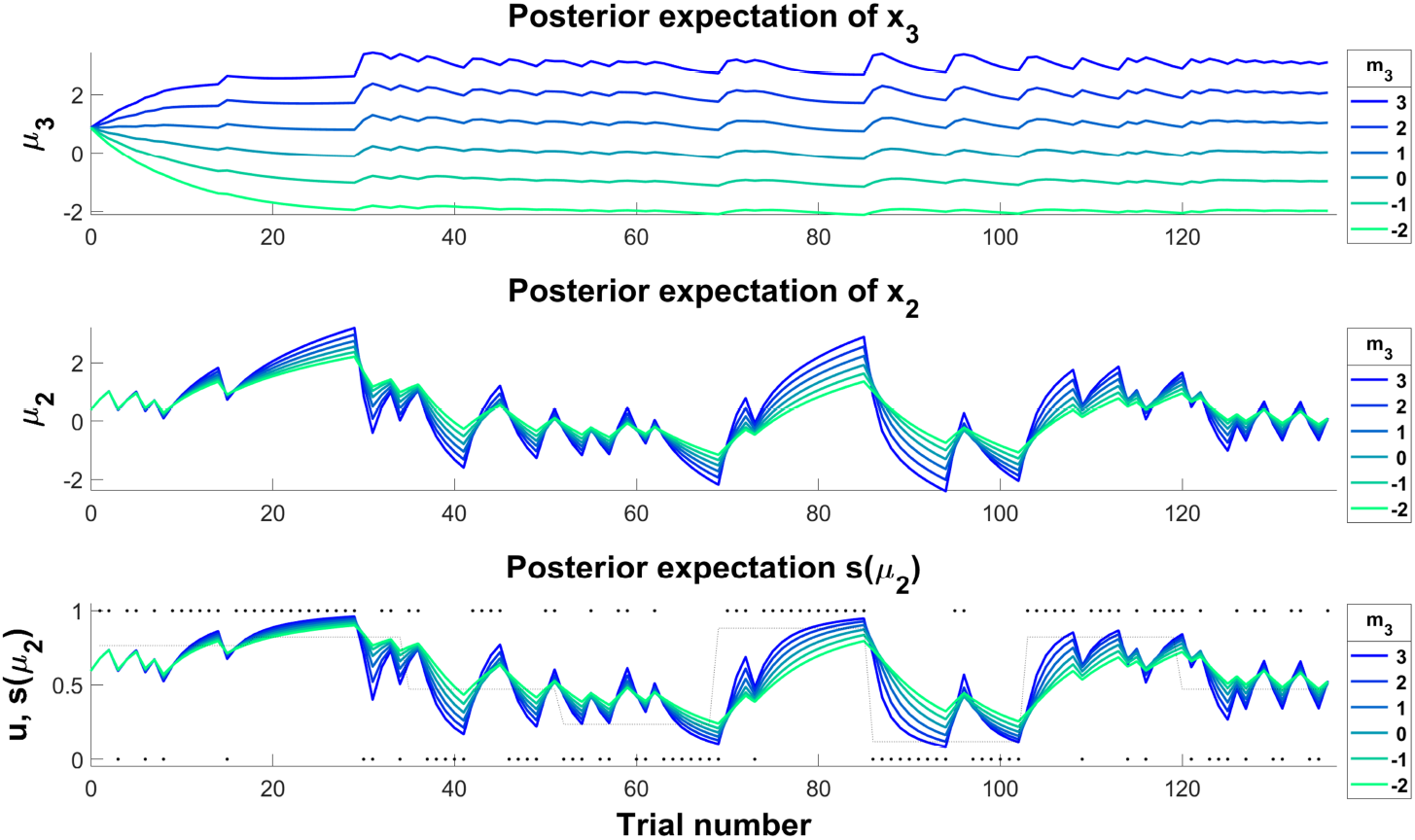
Simulating an altered perception of environmental volatility. Simulations showing the effect of changing the equilibrium point *m*_3_. Increasing *m*_3_ (colder colours) results in larger precision-weighted prediction errors leading to stronger belief updates across all levels of the hierarchy. Note, that high values of *m*_3_ also increase susceptibility to noisy inputs (e.g., trials 120-136). For the simulations, all other parameter values were fixed to the values of an ideal observer given the input.

##### Response model

The response model specifies how participants’ inference on the hidden states translates into decisions, i.e., to go with or against the advice. In our case the response model assumes that participants’ integrate the non-social cue *c*^(*k*)^ (the outcome probability indicated by the pie chart) and their belief that the adviser is providing accurate advice 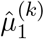 before seeing the outcome on the current trial *k*:

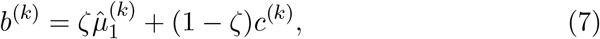

where *ζ* is a weight associated with the advice that expresses how much participants rely on the social information compared to the non-social cue.

The probability that a participant follows the advice (*y* = 1) can then be described by a sigmoid transformation of the integrated belief *b*:

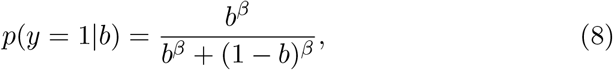

with

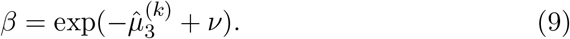

This relationship can be understood as a noisy mapping from the integrated beliefs to participants’ decisions, where the noise level is determined by the current prediction of the volatility of the advisers’ intentions 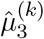, such that decisions become more deterministic (i.e., *exploitative*), if the environment is currently perceived as stable or more stochastic (i.e., *exploratory*), if the environment is perceived as volatile. Modelling the exploration-exploitation trade-off as a function of participants’ perception of volatility was favoured in previous model selection results using the same task (Diaconescu et al., 2014, 2017). Parameter *ν* is another subject-specific parameter that captures decision noise that is independent of the perception of volatility (lower values indicate larger decision noise). The prior mean and variance of this parameter was set based on previous studies that modelled learning about intentions (Diaconescu et al., 2020).

The models were implemented in Matlab (version: 2017a; https://mathworks.com) using the HGF toolbox (version: 3.0), which is made available as open-source code as part of the TAPAS (Frässle et al., 2021) software collection (https://github.com/translationalneuromodeling/tapas/releases/tag/v3.0.0). Perceptual models were implemented using the ‘tapas hgf binary’ function for the standard 3-level HGF and the ‘tapas hgf ar1 binary’ function for the mean-reverting HGF.

#### 2.5.2 Bayesian model selection

Based on our a simulation analysis (Diaconescu et al., 2019) and previous findings (Cole et al., 2020; Diaconescu et al., 2014, 2020; Reed et al., 2020), we formulated competing hypotheses about the computational mechanisms that could underlie emerging paranoid behaviour (Figure 3). A standard 3-level HGF (**Hypothesis I**) was compared to the mean-reverting HGF that assumed that learning about an adviser’s intentions was not only driven by hierarchical PE updates, but also included a drift process at the third level formalising the idea, that an altered perception of volatility underlies learning about others’ intentions in emerging psychosis (**Hypothesis II**; see also Figure 2). To arbitrate between the two hypotheses we performed random-effects Bayesian model selection (Rigoux et al., 2014; Stephan et al., 2009). Two additional control models were included, in which all parameters of the perceptual model were fixed to parameter values of an ideal Bayesian observer optimised based on the inputs alone using the ‘tapas bayes optimal binary’ function to assess whether perceptual model parameters needed to be estimated for either of the two main models. These “null” models assume that any variation in advice-taking behavior can be attributed solely to the response model parameters, i.e. the social bias and the decision noise.

**Figure 3:**
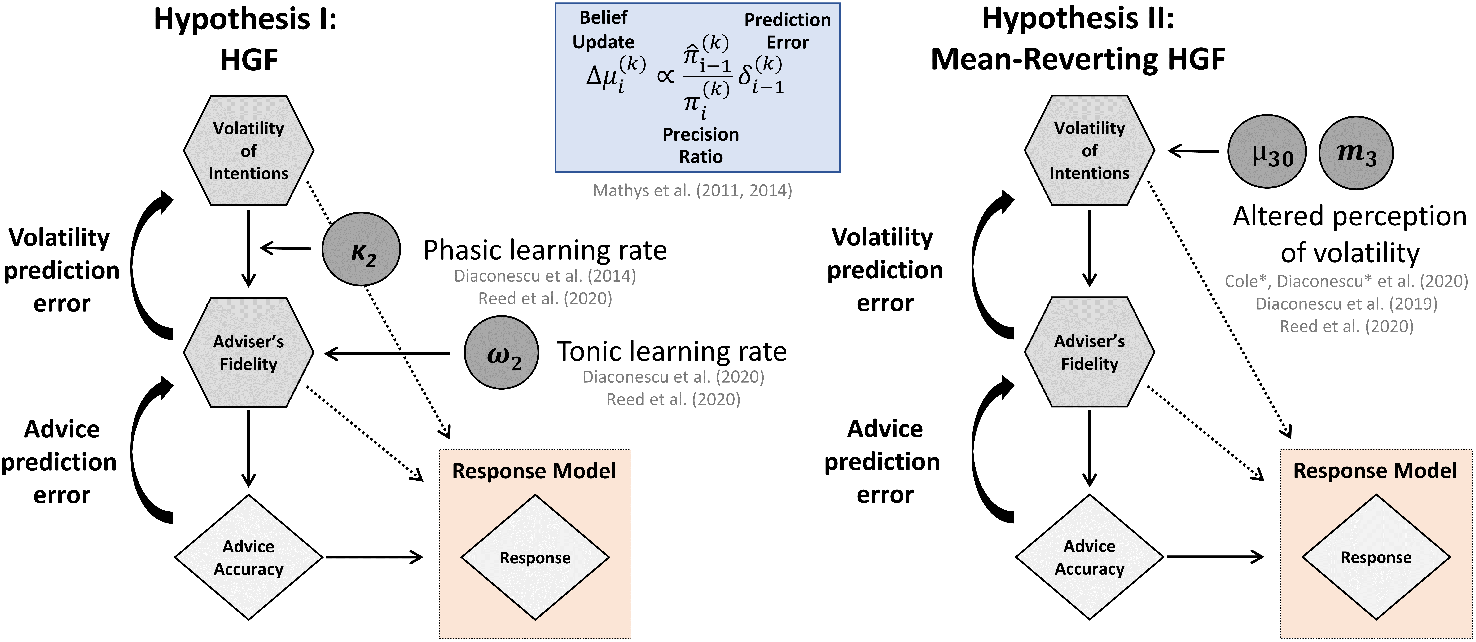
Model space. **Left**: Standard 3-level Hierarchical Gaussian Filter (HGF).(Mathys et al., 2011, 2014) **Right**: Mean-reverting HGF with a drift at the third level, which captures learning about the volatility of the adviser’s intentions. This model expresses the notion that early psychosis may be characterised by an altered perception of environmental volatility.

We report protected exceedance probabilities *ϕ*, which measure the probability that a model is more likely than any other model in the model space (Stephan et al., 2009), protected against the risk that differences between models arise due to chance alone (Rigoux et al., 2014). We also computed relative model frequencies *f* as a measure of effect size, which can be understood as the probability that a randomly sampled participant would be best explained by a given model. The model selection was implemented using the VBA toolbox (Daunizeau et al., 2014) (https://mbb-team.github.io/VBA-toolbox/).

#### 2.5.3 Model recovery

To assess whether models were recoverable, we conducted a series of simulations as done previously (Hauke et al., 2022). In brief, our model recovery analysis comprised simulating 20 synthetic datasets based on the empirical parameter estimates obtained from fitting all models to the empirical data of every participant. The sample size of each synthetic dataset was chosen to be equivalent to the empirical sample size (*N* = 56). The noise level was set based on the empirically estimated decision noise *ν*_*est*_. Each simulation was initialised using different random seeds to account for the stochasticity of the simulation. This led to a total of 4 (models) x 56 (participants) x 20 (simulation seeds) = 4,480 simulations. Subsequently, we re-inverted each of the proposed models on the synthetic data to determine, whether we could recover the true model under which synthetic data was generated. To assess model recovery, we then performed random-effects Bayesian model selection on each of the datasets with a sample size of *N* = 56 as in the empirical data and averaged the resulting protected exceedance probabilities across the 20 simulation seeds to obtain a model confusion matrix.

#### 2.5.4 Parameter recovery

In line with our previous work (Hauke et al., 2022), we also performed a parameter recovery analysis to determine whether model parameter estimates were reliable. Using the simulation and model inversion results from the model recovery analysis (see preceding section), we assessed how accurately the parameters generating the data (‘simulated’) corresponded to the parameters that were estimated when re-inverting the same model on that data (‘recovered’). We report Pearson correlations and their associated *p*-values to quantify our ability to recover the model parameters. Since, the significance of these correlations is influenced by sample size, we also computed Cohen’s *f* ^2^, where an *f* ^2^ ≥ 0.35 can be considered a large effect size (Cohen, 1988) and was interpreted as evidence for good parameter recovery.

### 2.6 Statistical analysis

We tested for differences in behaviour using a linear mixed-effects model with advice taking (#trials, in which participant went with the advice /# total trials) as the dependent variable and fixed effects for group and task phase (*stable* vs *volatile*), as well as a group-by-task-phase interaction as predictors of interest and age, working memory performance as covariates of no interest. Additionally, the model included a random intercept per participant.

Note, that including medication as a covariate is not recommended when comparing HC and patient groups. For completeness, however, we also report the results of mixed-effects model with current antipsychotic dose (100mg/day chlorpromazine equivalents) and current antidepressant dose (40mg/day fluoxetine equivalents) as covariates. Chlorpromazine equivalents were derived from The Maudsley^®^ prescribing guidelines in Psychiatry (Taylor et al., 2021) which is based on the literature and clinical consensus. Since paliperidone was not listed, equivalent estimates for paliperidone were based on Leucht et al. (2014). Fluoxetine equivalents were based on Hayasaka et al. (2015), with the exception of vorioxetin and citalopram which were not listed. For these, equivalents doses were assumed to be 10mg vortioxetin and 30mg citalopram, respectively, based on clinical practice.

Differences in model parameters were assessed using non-parametric Kruskal-Wallis tests. All statistical analyses were conducted in R (version: 4.04; https://www.r-project.org/) using R-Studio (version: 1.4.1106; https://www.rstudio.com/). We report both uncorrected *p*-values (*p*_*uncorr*_) and Bonferroni-corrected *p*-values adjusted for the number of free parameters (*n* = 7). Based on previous findings, we hypothesised that groups would differ with respect to coupling strength between the second and third level *κ*_2_ (Diaconescu et al., 2014; Reed et al., 2020), the evolution rate *ω*_2_ (Diaconescu et al., 2020; Reed et al., 2020), or parameters that are associated with the perception of volatility, i.e., the prior expectation about environmental volatility 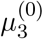 (Reed et al., 2020) or the equilibrium point of the drift at the third level *m*_3_ (Cole et al., 2020; Diaconescu et al., 2019).

## 3 Results

### 3.1 Sociodemographic and clinical characteristics

Sociodemographic and clinical characteristics are presented in Table 2.

**Table 2:**
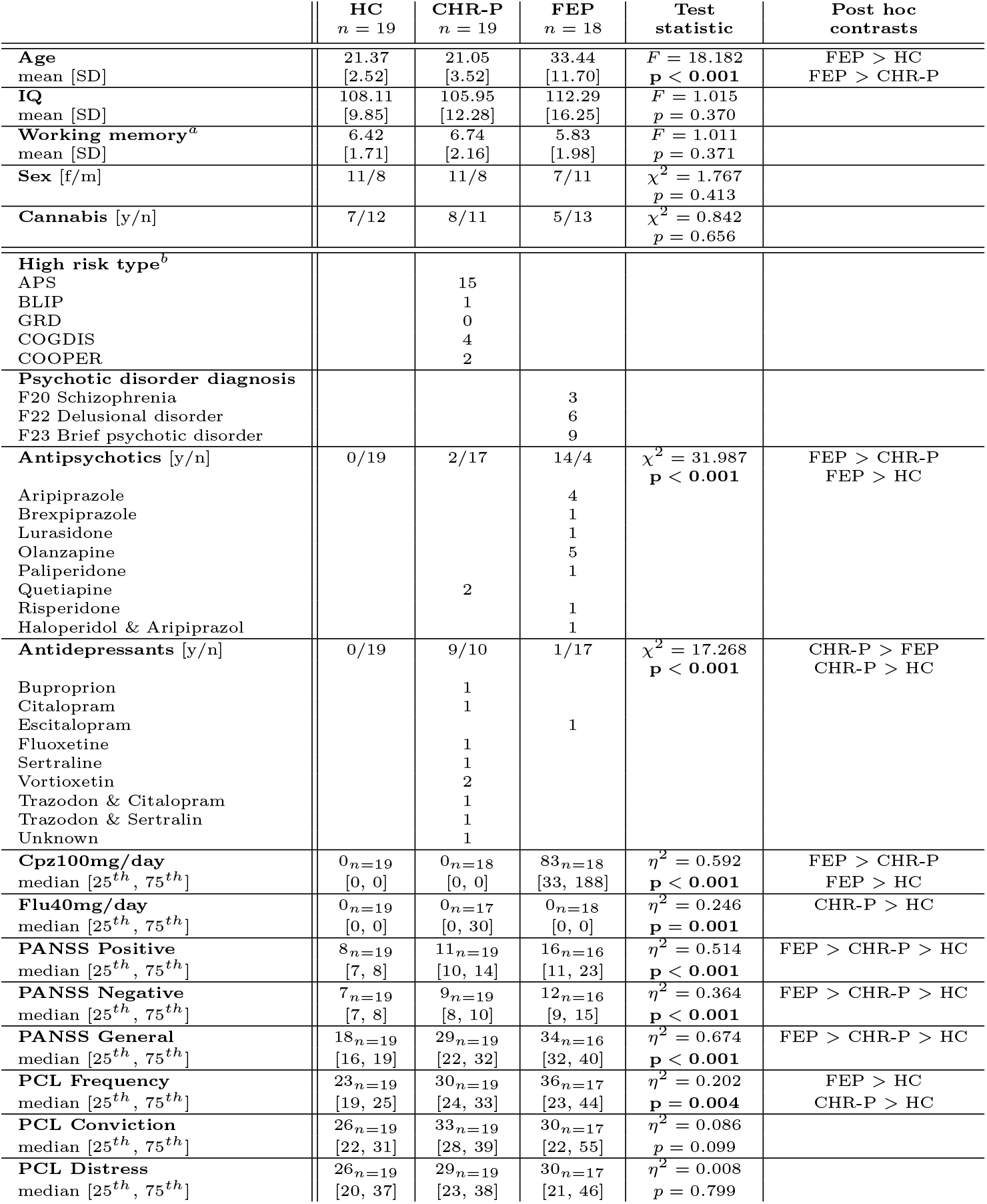
Demographic and clinical characteristics. All *p*-values are uncorrected. **HC**: Healthy controls. **CHR-P**: Individuals at clinical high risk for psychosis. **FEP**: First-episode psychosis patients. **APS**: Attenuated psychotic symptoms. **BLIP**: Brief and limited intermittent psychotic symptoms. **GRD**: Genetic risk and deterioration syndrome. **COGDIS**: Cognitive disturbances. **COPER**: Cognitive-perceptive basic symptoms. **Cpz100mg/day**: Antipsychotic equivalent dose for 100mg chlorpromazine per day. **Flu40mg/day**: Antidepressant equivalent dose for 40mg fluoxetine per day. **PANSS**: Positive and Negative Syndrome Scale.(Kay et al., 1987) **PCL**: Paranoia Checklist (Freeman et al., 2005). Bold print highlights *p*-values significant at: *p <* 0.05, uncorrected. ^**a**^ Assessed with the digit span backwards task from the Wechsler Adult Intelligence Scale–Revised (Wechsler, 1981). ^**b**^High risk types are not mutually exclusive.

|  | HC<br>*n* = 19 | CHR-P<br>*n* = 19 | FEP<br>*n* = 18 | Test<br>statistic | Post hoc<br>contrasts |
| --- | --- | --- | --- | --- | --- |
| **Age** | 21.37 | 21.05 | 33.44 | $F = 18.182$ | FEP > HC |
| mean [SD] | [2.52] | [3.52] | [11.70] | $p < \mathbf{0.001}$ | FEP > CHR-P |
| **IQ** | 108.11 | 105.95 | 112.29 | $F = 1.015$ | |
| mean [SD] | [9.85] | [12.28] | [16.25] | $p = 0.370$ | |
| **Working memory<sup>a</sup>** | 6.42 | 6.74 | 5.83 | $F = 1.011$ | |
| mean [SD] | [1.71] | [2.16] | [1.98] | $p = 0.371$ | |
| **Sex** [f/m] | 11/8 | 11/8 | 7/11 | $\chi^2 = 1.767$ | |
| | | | | $p = 0.413$ | |
| **Cannabis** [y/n] | 7/12 | 8/11 | 5/13 | $\chi^2 = 0.842$ | |
| | | | | $p = 0.656$ | |
| **High risk type<sup>b</sup>** |  |  |  |  |  |
| APS |  | 15 |  |  |  |
| BLIP |  | 1 |  |  |  |
| GRD |  | 0 |  |  |  |
| COGDIS |  | 4 |  |  |  |
| COOPER |  | 2 |  |  |  |
| **Psychotic disorder diagnosis** |  |  |  |  |  |
| F20 Schizophrenia |  |  | 3 |  |  |
| F22 Delusional disorder |  |  | 6 |  |  |
| F23 Brief psychotic disorder |  |  | 9 |  |  |
| **Antipsychotics** [y/n] | 0/19 | 2/17 | 14/4 | $\chi^2 = 31.987$ | FEP > CHR-P |
| | | | | $p < \mathbf{0.001}$ | FEP > HC |
| Aripiprazole |  |  | 4 |  |  |
| Brexpiprazole |  |  | 1 |  |  |
| Lurasidone |  |  | 1 |  |  |
| Olanzapine |  |  | 5 |  |  |
| Paliperidone |  |  | 1 |  |  |
| Quetiapine |  | 2 |  |  |  |
| Risperidone |  |  | 1 |  |  |
| Haloperidol & Aripiprazol |  |  | 1 |  |  |
| **Antidepressants** [y/n] | 0/19 | 9/10 | 1/17 | $\chi^2 = 17.268$ | CHR-P > FEP |
| | | | | $p < \mathbf{0.001}$ | CHR-P > HC |
| Bupropion |  | 1 |  |  |  |
| Citalopram |  | 1 |  |  |  |
| Escitalopram |  |  | 1 |  |  |
| Fluoxetine |  | 1 |  |  |  |
| Sertraline |  | 1 |  |  |  |
| Vortioxetin |  | 2 |  |  |  |
| Trazodon & Citalopram |  | 1 |  |  |  |
| Trazodon & Sertraline |  | 1 |  |  |  |
| Unknown |  | 1 |  |  |  |
| **Cpz100mg/day** | 0<sub>n=19</sub> | 0<sub>n=18</sub> | 83<sub>n=18</sub> | $\eta^2 = 0.592$ | FEP > CHR-P |
| median [25<sup>th</sup>, 75<sup>th</sup>] | [0, 0] | [0, 0] | [33, 188] | $p < \mathbf{0.001}$ | FEP > HC |
| **Flu40mg/day** | 0<sub>n=19</sub> | 0<sub>n=17</sub> | 0<sub>n=18</sub> | $\eta^2 = 0.246$ | CHR-P > HC |
| median [25<sup>th</sup>, 75<sup>th</sup>] | [0, 0] | [0, 30] | [0, 0] | $p = \mathbf{0.001}$ | |
| **PANSS Positive** | 8<sub>n=19</sub> | 11<sub>n=19</sub> | 16<sub>n=16</sub> | $\eta^2 = 0.514$ | FEP > CHR-P > HC |
| median [25<sup>th</sup>, 75<sup>th</sup>] | [7, 8] | [10, 14] | [11, 23] | $p < \mathbf{0.001}$ | |
| **PANSS Negative** | 7<sub>n=19</sub> | 9<sub>n=19</sub> | 12<sub>n=16</sub> | $\eta^2 = 0.364$ | FEP > CHR-P > HC |
| median [25<sup>th</sup>, 75<sup>th</sup>] | [7, 8] | [8, 10] | [9, 15] | $p < \mathbf{0.001}$ | |
| **PANSS General** | 18<sub>n=19</sub> | 29<sub>n=19</sub> | 34<sub>n=16</sub> | $\eta^2 = 0.674$ | FEP > CHR-P > HC |
| median [25<sup>th</sup>, 75<sup>th</sup>] | [16, 19] | [22, 32] | [32, 40] | $p < \mathbf{0.001}$ | |
| **PCL Frequency** | 23<sub>n=19</sub> | 30<sub>n=19</sub> | 36<sub>n=17</sub> | $\eta^2 = 0.202$ | FEP > HC |
| median [25<sup>th</sup>, 75<sup>th</sup>] | [19, 25] | [24, 33] | [23, 44] | $p = \mathbf{0.004}$ | CHR-P > HC |
| **PCL Conviction** | 26<sub>n=19</sub> | 33<sub>n=19</sub> | 30<sub>n=17</sub> | $\eta^2 = 0.086$ | |
| median [25<sup>th</sup>, 75<sup>th</sup>] | [22, 31] | [28, 39] | [22, 55] | $p = 0.099$ | |
| **PCL Distress** | 26<sub>n=19</sub> | 29<sub>n=19</sub> | 30<sub>n=17</sub> | $\eta^2 = 0.008$ | |
| median [25<sup>th</sup>, 75<sup>th</sup>] | [20, 37] | [23, 38] | [21, 46] | $p = 0.799$ | |

### 3.2 Behavioural results

We identified a significant group-by-task-phase interaction on the frequency of advice-taking (*F* = 5.275, *p* = 0.008; Figure 4A). To unpack this effect we repeated the analysis with three two-group models. We found significant group-by-task-phase interactions when comparing HC vs FEP (*F* = 8.520, *p*_*uncorr*_ = 0.006, *p* = 0.018 Bonferroni-corrected for the number of comparisons, i.e. *n* = 3) and HC vs CHR-P (*F* = 7.745, *p*_*uncorr*_ = 0.009, *p* = 0.026), but not when comparing CHR-P vs FEP (*F* = 0.047, *p*_*uncorr*_ = 0.830, *p* = 1.000), suggesting that both CHR-P and FEP showed reduced flexibility to take environmental volatility into account as the difference between stable and volatile phase was reduced compared to HC. None of the covariates significantly impacted advice taking.

**Figure 4:**
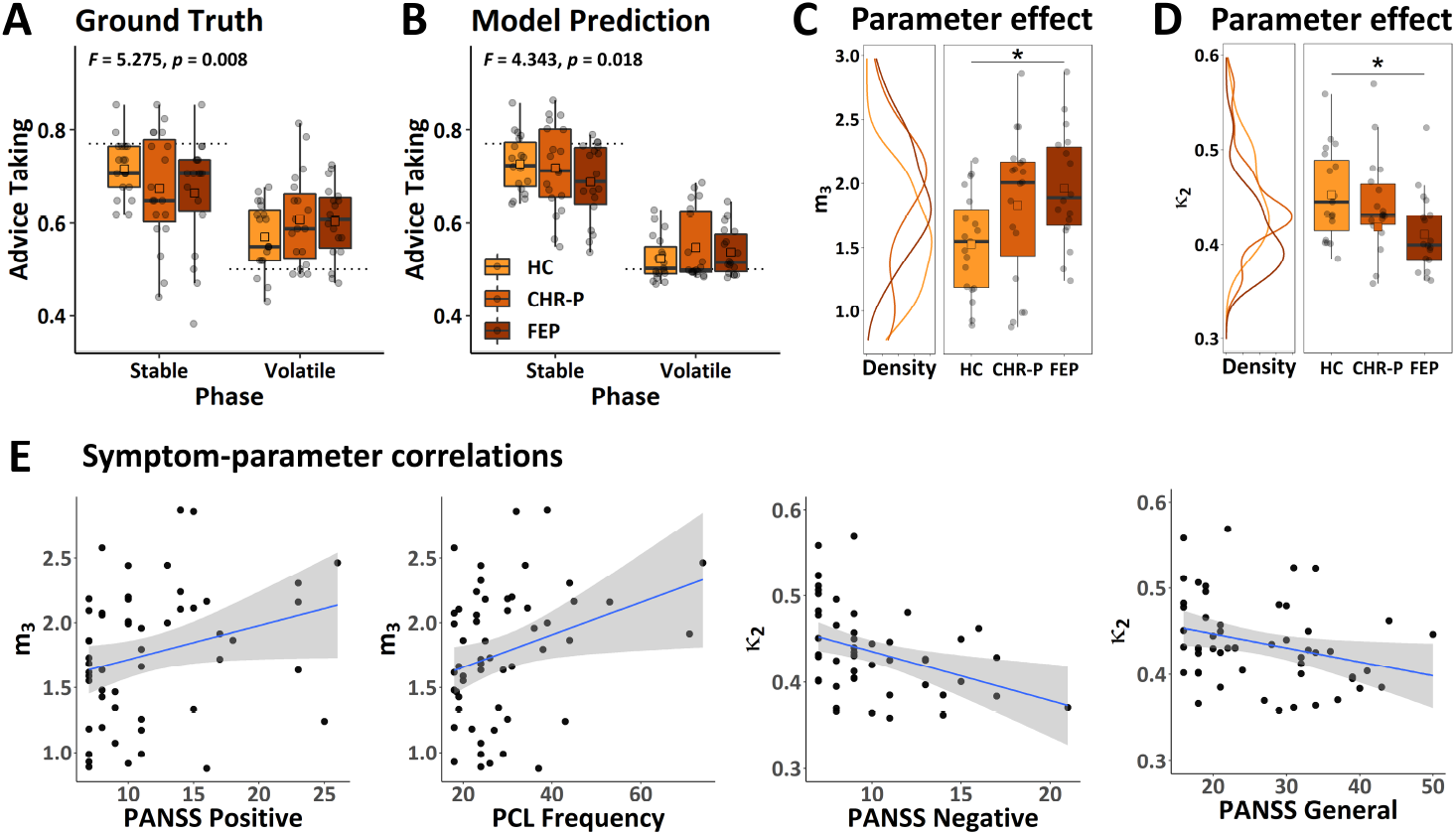
Behavioural results and parameter group effects. **A** Behavioural results (ground truth). Black dashed lines indicate the average accuracy of advice for each of the two phases. **B** Model prediction. **C** Parameter effect for drift equilibrium point *m*_3_. **D** Parameter effect for coupling strength *κ*_2_. **E** Correlation between model parameters and either Positive and Negative Syndrome Scale (Kay et al., 1987) (**PANSS**) or Paranoia Checklist (Freeman et al., 2005) (**PCL**). Note, that raw scores are displayed for illustration purposes only. Statistical analyses were conducted using nonparametric Kendall rank correlations. Displayed regression lines were computed using a linear model based on the raw scores. Note, that one outlier (*κ*_2_ = 0.006) was removed for displaying the effect on *κ*_2_ in **D** and **E**. This outlier was outside of 7× the interquartile range. Excluding this participant did not affect the significance of the results. **P**: Positive symptoms. **N**: Negative symptoms. **G**: General symptoms. *F* - and *p*-values indicate results of ANCOVAs corrected for working memory performance, antipsychotic medication, antidepressant medication, and age. Boxes span the 25^*th*^ to 75^*th*^ quartiles and whiskers extend from hinges to the largest and smallest value that lies within 1.5× interquartile range. Asterisks indicate significance of non-parametric Kruskal-Wallis tests at: **\*** *p <* 0.05, using Bonferroni correction.

The group-by-task-phase interaction remained significant after including antipsychotic and antidepressant dose as covariates (*F* = 4.900, *p* = 0.011). Neither the effect of antipsychotic dose (*F* = 0.006, *p* = 0.939) or antidepressant dose (*F* = 0.112, *p* = 0.739) were significant. Unpacking this model again revealed significant group-by-task-phase interactions when comparing HC vs FEP (*F* = 8.520, *p*_*uncorr*_ = 0.006, *p* = 0.018), but not when comparing CHR-P vs FEP (*F* = 0.671, *p*_*uncorr*_ = 0.419, *p* = 1.00). The group-by-task-phase interaction effect in HC vs CHR-P did not survive Bonferroni correction (*F* = 5.154, *p*_*uncorr*_ = 0.030, *p* = 0.089).

### 3.3 Modelling results

#### 3.3.1 Bayesian model selection and model recovery

The model recovery analysis (Figure 6) indicated that the control models (CI and CII) could not be well-distinguished. This was likely due to the fact that the equilibrium point *m*_3_ in CII was optimised based on the input alone, which resulted in a value for *m*_3_ that was close to the prior, rendering the predictions of the two control models very similar. Most importantly, however, the two main models associated with Hypothesis I and II could be well-distinguished.

**Figure 6:**
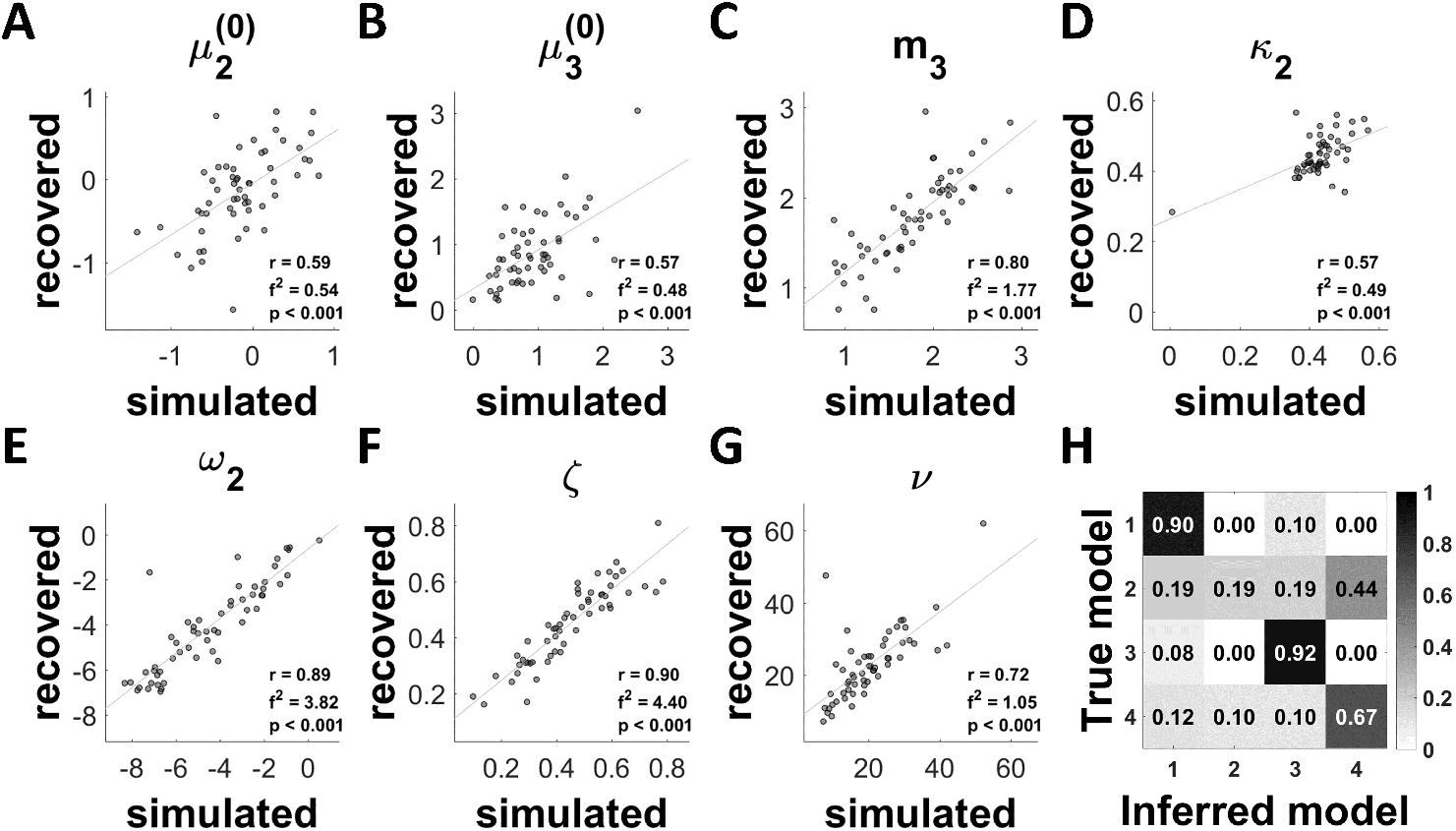
Model and parameter recovery analyses. **A-G** Parameter recovery result for one random seed for the mean-reverting HGF with drift at the 3^*rd*^ level (Hypothesis II; Figure 3). **H** Model recovery analysis. The grey scale indicates protected exceedance probability averaged across all 20 random seeds.

After confirming that the two hypotheses were distinguishable, we first performed Bayesian model selection including participants from all groups. The results were inconclusive (*ϕ* = 74.03%, *f* = 53.80% in favour of Hypothesis II) possibly suggesting that different groups were best explained by different models (i.e., different computational mechanisms). To assess this possibility, we repeated the model selection for each group separately (Figure 5A). In HC, the winning model was the standard 3-level HGF (Hypothesis I; *ϕ* = 96.63%, *f* = 95.93%). Conversely, in FEP the mean-reverting HGF that included a drift at the third level was selected (Hypothesis II; *ϕ* = 99.95%, *f* = 95.92%). For CHR-P, we observed a more heterogeneous results: While the mean-reverting model was favoured (Hypothesis II; *ϕ* = 84.50%, *f* = 60.24%), there was also evidence for the standard HGF, albeit to a much lesser extent (Hypothesis I; *ϕ* = 14.41%, *f* = 37.19%). Further inspection of the model attributions for all individual participants revealed an interesting pattern (Figure 5B). All HC were attributed to the standard HGF with over 97% probability, whereas FEP were attributed to the mean-reverting model with over 99%. Interestingly, model attributions for CHR-P were more heterogeneous ranging from 0 to 100% probability, suggesting that some individuals were better explained by the standard HGF, but others by the mean-reverting model.

**Figure 5:**
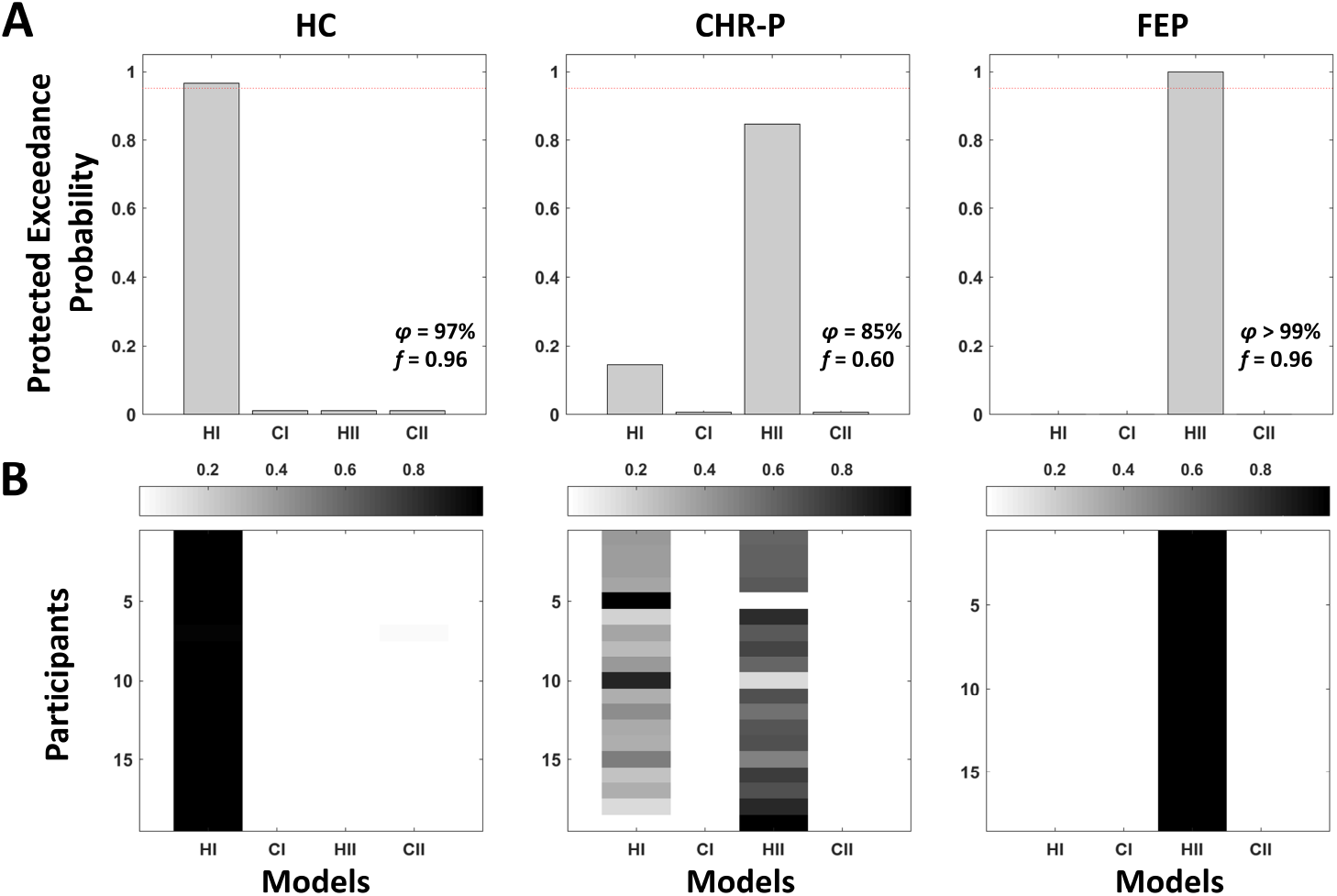
Bayesian model selection results. **A** Protected exceedance probabilities for within-group random-effects Bayesian model selection(Stephan et al., 2009; Rigoux et al., 2014) to arbitrate between Hypothesis I (**HI**; standard 3-level HGF) and Hypothesis II (**HII**; mean-reverting HGF with drift at 3^*rd*^ level in line with an altered perception of volatility). Two corresponding control models were included (**CI** and **CII**), for which the perceptual model parameters were fixed. Model selection was performed separately in healthy controls (**HC**), individuals at clinical high risk for psychosis (**CHR-P**), or first-episode psychosis patients (**FEP**). The dashed line indicates 95% exceedance probability. **B** Model attributions for each participant.

#### 3.3.2 Posterior predictive checks and parameter recovery

To assess whether the mean-reverting model (Hypothesis II) captured the behavioural effects of interest, we conducted posterior predictive checks by repeating the behavioural analysis on this model’s predictions. This analysis confirmed that the mean-reverting model recapitulated the group-by-task-phase interaction effect on advice-taking frequency (*F* = 4.343, *p* = 0.018; Figure 4B). We also repeated all three two-group models on the model predictions and found a significant group-by-task-phase interaction when comparing HC vs FEP (*F* = 8.337, *p*_*uncorr*_ = 0.007, *p* = 0.020) and no significant interaction when comparing CHR-P vs FEP (*F* = 1.106, *p*_*uncorr*_ = 0.300, *p* = 0.900) as before in the empirical data. The group-by-task-phase interaction did not reach significance for HC vs CHR-P (*F* = 3.662, *p*_*uncorr*_ = 0.064, *p* = 0.191).

Our parameter analysis indicated good recovery (i.e., Cohen’s *f* ^2^ ≥ 0.35) for four out of the seven model parameters including the drift equilibrium point *m*_3_ (Figure 6). However, recovery for 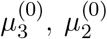, and *κ*_2_ fulfilled this criterion only in 55%, 65%, and 55% of the simulations respectively.

When inspecting parameter identifiability, we observed unconcerning correlations between all pairs of parameters (*r* ≤ |0.39|) except for the correlation between *m*_3_ and *ν* (*r* = 0.93). To assess, whether we could address this colinearity by removing one of the parameters from the response model, we formulated two alternative model families in which either decision noise parameter *ν* or state 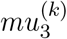 was removed from the response model and compared them to the original model family (cf. Eq. 9). These alternative model families assumed either that the estimated volatility was solely driving the mapping of beliefs to decisions (response model family two without decision noise parameter) or the decision noise alone determined belief-to-response mapping (response model family three that excluded the estimated volatility from the response model). Family-level inference (Penny et al., 2010) indicated that the original model family that included both state 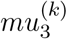 and parameter *ν* was the winning family compared to family two without 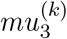 (exceedance probability: 100.00%, *f* = 99.12%) and family three without *ν* (exceedance probability: 99.54%, *f* = 66.66%). We thus concluded that both 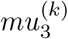 and *ν* should be included in the response model as they reflect two important mechanisms determining the exploration-exploitation trade-off. First, the estimated volatility captures the impact of learning on belief-to-response mapping, i.e., more exploration when the adviser’s intentions are perceived as volatile and more exploitation when the adviser’s intentions are perceived to be stable. Second, the decision noise captures non-inference related sources of noise, for example, due to distractions or incorrect button presses. However, the interpretation of *m*_3_-effects reported below should be taken as preliminary and needs to be confirmed in another study with a volatility schedule that is optimised for decorrelating of these parameters.

#### 3.3.3 Parameter group effects

The model selection indicated that the mean-reverting model was a better explanation for behaviour of FEP, but not of HC. In this situation, it is generally recommended to investigate parameter group effects using Bayesian model averaging (Stephan et al., 2010). However, we were interested in assessing *why* this model was selected for FEP. Specifically, we wanted to investigate whether the perception of volatility in FEP increased or decreased over time (see also simulations illustrating these two possibilities in Figure 2), because our a priori hypothesis was that individuals with emerging psychosis should perceive the environment as increasingly volatile (increased *m*_3_ compared to controls; Diaconescu et al. (2019)). To distinguish between these two possibilities, we compared the drift equilibrium point *m*_3_ across the three groups and found that *m*_3_ was significantly different across the groups (*η*^2^ = 0.142, *p*_*uncorr*_ = 0.020). Post hoc tests revealed that *m*_3_ was significantly increased in FEP compared to HC suggesting that FEP perceived the intentions of the adviser as increasingly more volatile over time (*η*^2^ = 0.212, *p* = 0.017, Bonferroni-corrected for the number of comparisons across groups, i.e., *n* = 3; Figure 4C). We also performed an exploratory analysis including all other free model parameters. This analysis revealed an additional effect on coupling strength *κ*_2_ (*η*^2^ = 0.138, *p*_*uncorr*_ = 0.022), which was driven by reduced coupling strength between the second and third level of the perceptual hierarchy in FEP compared to HC (*η*^2^ = 0.217, *p* = 0.016, Bonferroni-corrected for the number of comparisons across groups, i.e., *n* = 3; Figure 4D). However, neither the effect on *m*_3_ nor *κ*_2_ survived Bonferroni correction for the number of parameters, i.e. *n* = 7 (*p* = 0.140 and *p* = 0.157, respectively).

#### 3.3.4 Symptom-parameter correlations

Some authors (e.g., Esterberg and Compton (2009)) have argued that psychosis may be better conceptualised on a continuum rather than categorically, based on evidence that a significant percentage of the general populations reports some psychosis symptoms (Kendler et al., 1996; Tien, 1991). In line with this proposal, we assumed a continuum perspective and investigated whether the equilibrium point *m*_3_ and coupling strength *κ*_2_ were correlated with specific symptom subscales of the Positive and Negative Syndrome Scale (PANSS) (Kay et al., 1987) across all three groups with non-parametric Kendall rank correlations (see Figure 4E).

We found a positive correlation between *m*_3_ and PANSS positive symptoms (*τ* = 0.203, *p*_*uncorr*_ = 0.038) and negative correlations between *κ*_2_ and PANSS negative and general symptoms (*τ* = −0.253, *p*_*uncorr*_ = 0.011 and *τ* = −0.219, *p*_*uncorr*_ = 0.022 respectively). Firstly, this suggest that individuals who perceived the adviser’s intentions to be increasingly volatile over time also experienced more severe positive psychosis symptoms. Secondly, the negative correlation between *κ*_2_ and PANSS negative and general symptoms implies that individuals with more severe negative and general symptoms displayed lower *κ*_2_ values or a decoupling between the third and the second levels of the hierarchy. These correlations, however, did not survive Bonferroni correction (*p* = 0.228, *p* = 0.068, and *p* = 0.132 respectively, adjusted for 2 (#parameters) x 3 (#PANSS subscales) = 6 comparisons).

Since the PANSS (Kay et al., 1987) was specifically designed to assess symptom expression in clinical populations, we also calculated correlations with the Paranoia Checklist (PCL) (Freeman et al., 2005), an instrument more sensitive to expressions of paranoia in healthy or subclinical populations. We found a correlation between *m*_3_ and the PCL frequency subscale (*τ* = 0.201, *p*_*uncorr*_ = 0.034), indicating that individuals who perceived the adviser’s intentions to be increasingly volatile over time also reported a higher frequency of paranoid beliefs. Again, this correlation did not survive Bonferroni correction (*p* = 0.204, adjusted for 2 (#parameters) x 3 (#PCL subscales) = 6 comparisons).

## 4 Discussion

In this study, we investigated the computational mechanisms underlying emerging psychosis. Our model selection results suggest that FEP may operate under a different computational mechanism compared to HC that is characterised by perceiving the environment as *increasingly volatile*. A strength of our study is that this effect is unlikely due to long term medication effects as FEP were only briefly medicated. Furthermore, we observed more heterogeneity in CHR-P, possibly indicating that this modelling approach may be useful to stratify the CHR-P population and identify individuals that are more likely to transition to psychosis. Assuming a psychosis continuum perspective, we also found tentative evidence suggesting that the drift equilibrium point *m*_3_ and the coupling strength between hierarchical levels *κ*_2_ may be affected in emerging psychosis and that these parameters provide a clinically relevant description of individuals’ learning profiles. However, due to the small sample size, these results should be interpreted with caution.

### 4.1 Related modelling work

Bayesian accounts of psychosis (Fletcher and Frith, 2009; Sterzer et al., 2018; Adams et al., 2022) propose that psychosis may be characterised by aberrant PEs that provide the breeding ground for delusions to form. Our results are in line with these proposals and the predictions of increased precision-weighted PE-learning in early psychosis derived through simulations (Diaconescu et al., 2019). Moreover, our results enable a more nuanced characterisation and point towards an altered perception of environmental volatility as a possible consequence of aberrant PE learning. Specifically, perceiving the intentions of another person as increasingly volatile over time translates to reduced precision of beliefs about environmental volatility. This, in turn, results in larger precision-weighted PEs through decreasing the denominator of the precision ratio that weighs PEs (see Equation 5). This finding is in line with Bayesian accounts, although we cannot say whether changes in the perception of volatility are caused by aberrant PEs or vice versa without longitudinal assessment of changes within the same participants. However, we note that the mean-reverting model was only conclusively selected in the FEP group and not already in the CHR-P group, although the mean-reverting model was favoured in the model attributions for some CHR-P individuals (Figure 5B). In contrast to our a priori hypothesis (Diaconescu et al., 2019), we did not find evidence for a compensatory increase in the precision of high-level priors or reduced learning (e.g., reduced evolution rate *ω*_2_) in patients who have strong conviction in their delusional beliefs. This was also proposed as a cognitive mechanism to make sense of aberrant PEs by Kapur (2003) and observed empirically by others in healthy participants with paranoid ideations (Diaconescu et al., 2020; Wellstein et al., 2020) as well as patients with schizophrenia, (Baker et al., 2019), although Baker et al. (2019) used a non-social probabilistic reasoning task.

Reed et al. (2020) employed the HGF to investigate the computational mechanisms underlying paranoia in a subclinical population and schizophrenia patients using a non-social reversal learning task. They found increased expected volatility 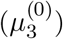 in participants with higher levels of paranoia using the standard 3-level HGF. Our model selection suggested that this model explains behaviour better in HC, whereas FEP were better characterised by a mean-reverting HGF that included a drift at the third level. It should be noted that increasing 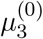 and including a drift at the third level, which increases over time, can both be interpreted as expecting the environment to be more volatile, but the drift provides a more nuanced description of changes that occur *during* the learning session. Our results are thus in line with previous results, but possibly provide a perspective that takes within-task dynamics more explicitly into account (see simulations in the Supplement). An interesting observation based on simulations is that artificial agents with increased *m*_3_ are quicker to adapt to volatile changes between very helpful and very misleading advice (trials 68-119), but increasing *m*_3_ also leads to more susceptibility to noisy inputs following this period of rapid, but meaningful changes (trials 120-136; Supplement).

Moreover and in contrast to our results, Reed et al. (2020) found increased and not reduced coupling strength *κ*_2_. This discrepancy may be related to differences in the tasks employed (non-social three-option reversal learning task vs our social learning task), but we also note that *κ*_2_ was not always well-recoverable in our simulation analysis. Therefore, we do not wish to draw strong conclusions based on the *κ*_2_ effect in our study, although we found effects suggesting that *κ*_2_ may be related to negative and general symptoms.

### 4.2 Is the perception of environmental volatility altered specifically in social contexts?

Here, we employed an ecologically valid social learning task (Diaconescu et al., 2014, 2017) to study changes in learning about other’s intentions. Some authors (Reed et al., 2020; Suthaharan et al., 2021) have raised the question of whether changes in learning like the ones observed in this study are reflective of a specifically social or rather a domain-general learning deficit. Here, we did not assess whether differences with respect to the perception of environmental volatility were specific to a social context since we did not include a non-social control task. However, it will be important to address this question in future studies.

Interestingly, recent studies also identified a mean-reverting HGF with a drift towards larger volatility estimates as the winning model in a sample of CHR-P participants (Cole et al., 2020) and changes in *m*_3_ to be associated with a schizophrenia diagnosis (Fromm et al., 2022) in non-social, two-option reversal learning tasks. Others found changes in model parameters related to the perception of environmental volatility in healthy, subclinical, and schizophrenia patient populations (Reed et al., 2020; Suthaharan et al., 2021). Reed et al. (2020) also included a social control task, which did not affect the parameter effects. Therefore, this mechanism may not be specifically tied to social contexts, but instead may be related to a more general deficit in learning under uncertainty (Reed et al., 2020; Suthaharan et al., 2021). However, we do note that the social control task employed by Suthaharan et al. (2021) was not as ecologically valid as other tasks that were used to study paranoia such as the dictator game (Raihani and Bell, 2017; Barnby et al., 2020, 2022) or our task which was adapted from empirically-observed human-human interactions in a previous study using videos of human advisers intending to either help or deceive players (Diaconescu et al., 2014). Finally, it is also possible that there are both domain-general and domain-specific changes, but that these can only be studied at the neuronal level and converge on the same behavioural model parameters.

### 4.3 What causes an altered perception of volatility?

Interestingly, there may be at least two possibly interacting pathways that can lead to an altered perception of environmental volatility. First, abnormalities in monoamine systems may lead to aberrant PEs that are unpredictable and lead to the expectation that the environment is very volatile (Diaconescu et al., 2019; Kapur, 2003). In line with this pathway, Reed et al. (2020) found that methamphetamine administration induced changes in model parameters that impacted learning about environmental volatility in rats. Moreover, Diaconescu et al. (2017) found activation in dopaminoceptive regions such as the dopaminergic midbrain during the same social learning task that was used in the current study. Similarly, unstable dynamics in cortical circuits (related to synaptic dysfunction, or indeed abnormal neuromodulation) may also increase updating to unexpected evidence and thus increase the perception of environmental volatility (Adams et al., 2018; Hauke et al., 2022). Secondly, external shifts in the volatility of the environment, like for example the global health crisis of the COVID-19 pandemic, may also result in an altered perception of volatility and emergence of paranoid thoughts or endorsement of conspiracy theories (Suthaharan et al., 2021). This second (environmental) pathway may also be relevant for understanding increased incidence of schizophrenia in individuals that experience migration (Selten et al., 2020) and those living in urban environments (Vassos et al., 2012) as individuals exposed to both of these risk factors may be confronted with – in some cases drastically – changing environments. In summary, there are likely multiple possibly interacting pathways that could give rise to an altered perception of environmental volatility.

### 4.4 Clinical implications

We identified trend-correlations between the drift equilibrium point *m*_3_ and PANSS positive symptoms and the frequency of paranoid thoughts and between the coupling strength *κ*_2_ and PANSS negative and general symptoms. While the evidence was not conclusive in this study since these correlations were not significant after multiple testing correction, we note that the effects were in the expected direction, such that perceiving the environment as increasingly volatile (higher *m*_3_) was associated with higher frequency of paranoid thoughts and more severe positive symptoms in general. Additionally, increased decoupling of the third level from the second level of the HGF, which leads to aberrant learning under uncertainty, correlated with more severe negative symptoms. Future well-powered studies are needed to assess whether these effects can be confirmed in larger samples. Interestingly, we observed heterogeneous model attributions specifically in CHR-P, whereas the model selection clearly favoured the standard 3-level HGF in HC and the mean-reverting model in FEP. This finding suggests that this model may be helpful to identify CHR-P patients that will more likely transition to a psychotic disorder.

### 4.5 Limitations

Several limitations of this study merit attention. First, the sample size of this study was small due to very selective inclusion criteria with respect to medication, which, however, enabled us to minimise the impact of long term medication effects. Larger studies with a volatility structure optimised to decorrelate *m*_3_ and *ν* are needed to replicate our results and increase statistical power to identify correlations between model parameters and symptoms.

Secondly, we cannot assess the specificity of our results with respect to the social domain since we did not include a non-social control task. Lastly, we also cannot speak to the specificity with respect to other diagnoses, because we did not include a clinical control group, which is an important avenue for future research.

### 4.6 Future directions

While we found evidence for increased uncertainty associated with higher-level beliefs about the volatility of others’ intentions, future studies will have to examine whether a compensatory increase in the precision of higher-level beliefs occurs during later stages of schizophrenia, possibly also fluctuating with the severity of psychosis, or whether other models are better suited to capture the conviction associated with delusory beliefs during acute psychotic states (e.g., Baker et al. (2019); Erdmann and Mathys (2021); Adams et al. (2022)). Furthermore, the neural correlates of belief updating in emerging psychosis during social learning should be examined to identify neural pathways that may underlie the changes in perception that were suggested by the model. Lastly, longitudinal studies are needed to assess whether model parameters can be leveraged as predictors for transition to psychosis or treatment response in individual patients with psychosis.

### 4.7 Conclusions

In conclusion, our results suggest that emerging psychosis is characterised by an altered perception of environmental volatility. Furthermore, we observed heterogeneity in model attributions in individuals at high risk for psychosis suggesting that this computational approach may be useful to stratify the high risk state and for predicting transition to psychosis in clinical high risk populations.

## Data Availability

De-identified data will be made available under https://osf.io/ upon acceptance of this manuscript. Note, that one participant did not consent to make their data available for reuse and will be excluded from the public repository. To ensure reproducibility, we report all results excluding this participant in the Supplement.

## Data Accessibility

The analysis code for this study is publicly available under https://github.com/Murdugan/compi_ioio_phase. Data will be made available under https://osf.io/upon acceptance of this manuscript. Note, that one participant did not consent to make their data available for reuse and was excluded from the public repository. To ensure reproducibility, we report all results excluding this participant in the Supplement.

## Acknowledgements

We thank the participants for volunteering their energy and valuable time despite all the challenges they faced allowing us to pursue this research and the Schizophrenia International Research Society for honoring this work with the best poster price at the 2022 Congress of the Schizophrenia International Research Society. Furthermore, we also acknowledge that previous version of this article was published as part of DJH’s PhD thesis (Hauke, 2022) and made available as a preprint on https://www.medrxiv.org/.

## Authors’ Contributions

DJH had full access to all of the data in the study and takes responsibility for the integrity of the data and the accuracy of the data analysis. All authors contributed substantially to this work as outlined below in alphabetical order:

*Concept and design*: AOD, DJH, SB

*Acquisition, analysis, or interpretation of data* : All authors.

*Drafting of the manuscript* : DJH

*Critical revision of the manuscript for important intellectual content* : All authors.

*Obtained funding*: AOD, DJH

*Administrative, technical, or material support* : AOD, CA, SB, VR

*Supervision*: AOD, VR

