## supplement for "Aberrant perception of environmental volatility during social learning in emerging psychosis"

### 1 Simulations

Supplementary Figures 1 and 2 highlight the different impact that changes in the drift equilibrium point $m_3$ and changes in the prior expectation about environmental volatility $\mu_3^{(0)}$ have on belief trajectories at different levels of the inferential hierarchy and on simulated behavior. For completeness, we also included simulations illustrating the effects of changing the coupling between hierarchical levels $\kappa_2$ (Supplementary Figure 3) and changing the evolution rate $\omega_2$ (Supplementary Figure 4).

#### 1.1 Simulating changes in the equilibrium point

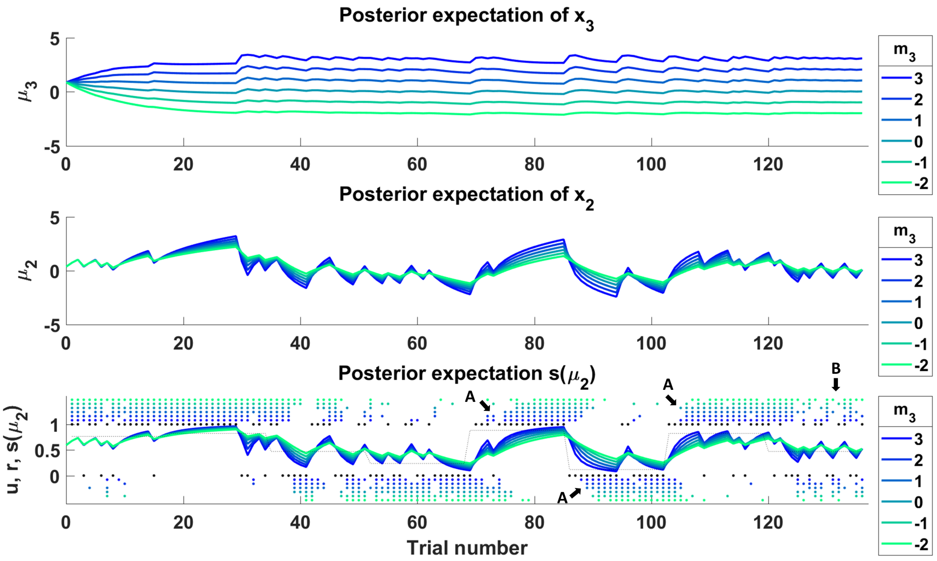

Supplementary Figure 1: **Simulations showing the effect of changing the equilibrium point** $m_3$. Shown are trajectories of beliefs about the volatility the adviser's intentions $\mu_3$ (upper panel), beliefs about the adviser's fidelity $\mu_2$ (middle panel) and about the advice accuracy $s(\mu_2)$. Black dots indicate inputs (1: helpful advice, 0: misleading advice) and colored dots simulated responses (1: going with the advice, 0: going against the advice). Increasing $m_3$ (colder colours) results in larger precision-weighted prediction errors leading to stronger belief updates across all levels of the hierarchy that increase over the course of the session. The effect on behavior depends on the input structure. When agents are exposed to volatile changes between very helpful and very misleading advice (trials 68-119), higher $m_3$ leads agents to detect changes more rapidly (see black arrows labelled A). However, high values of $m_3$ also increase susceptibility to noisy inputs (e.g., trials 120-136; see black arrow labelled B). For the simulations, all other parameter values were fixed to the values of an ideal observer given the input.

#### 1.2 Simulating changes in the prior expectation about environmental volatility

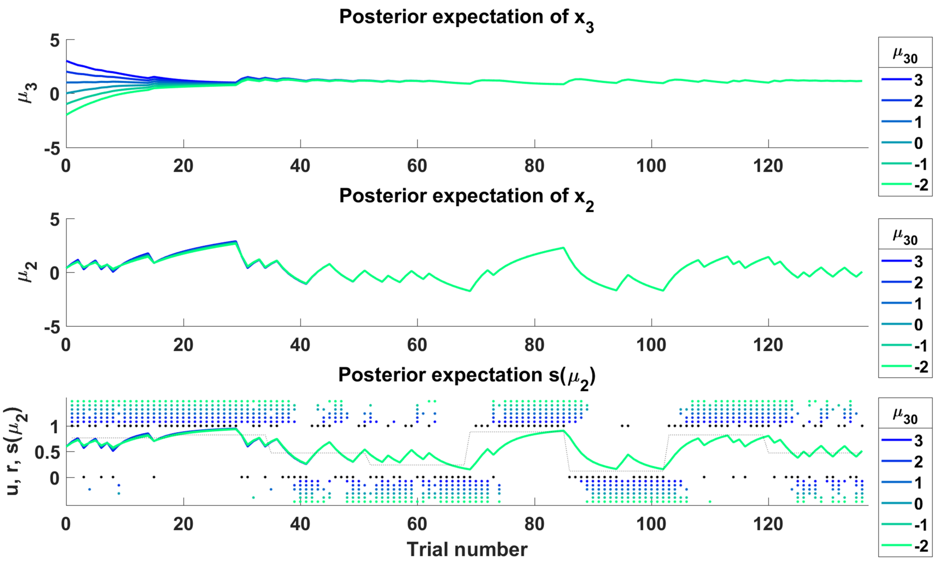

Supplementary Figure 2: **Simulations showing the effect of changing the prior expectation about environmental volatility** $\mu_3^{(0)}$. Shown are trajectories of beliefs about the volatility the adviser's intentions $\mu_3$ (upper panel), beliefs about the adviser's fidelity $\mu_2$ (middle panel) and about the advice accuracy $s(\mu_2)$. Black dots indicate inputs (1: helpful advice, 0: misleading advice) and colored dots simulated responses (1: going with the advice, 0: going against the advice). Increasing $\mu_3^{(0)}$ (colder colours) results in changes primarily in the first trials of the session and changes do not propagate strongly to lower levels.

#### 1.3 Simulating changes in the coupling strength

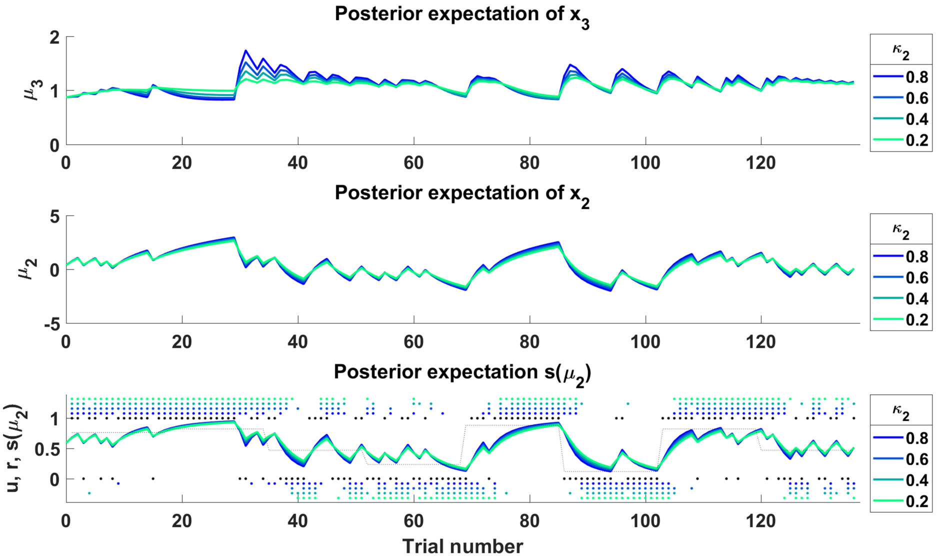

Supplementary Figure 3: **Simulations showing the effect of changing the coupling strength** $\kappa_2$. Shown are trajectories of beliefs about the volatility the adviser's intentions $\mu_3$ (upper panel), beliefs about the adviser's fidelity $\mu_2$ (middle panel) and about the advice accuracy $s(\mu_2)$. Black dots indicate inputs (1: helpful advice, 0: misleading advice) and colored dots simulated responses (1: going with the advice, 0: going against the advice).

#### 1.4 Simulating changes in the evolution rate

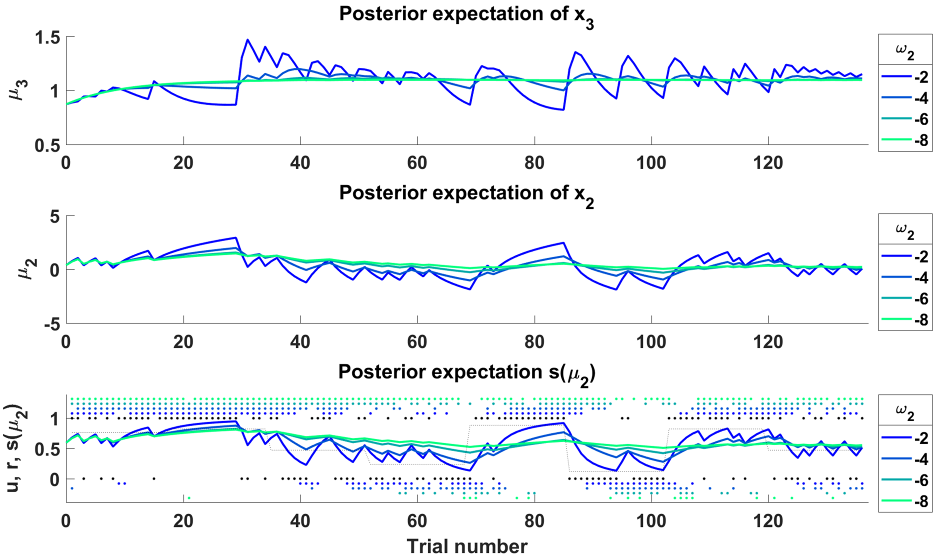

Supplementary Figure 4: **Simulations showing the effect of changing the evolution rate** $\omega_2$. Shown are trajectories of beliefs about the volatility the adviser's intentions $\mu_3$ (upper panel), beliefs about the adviser's fidelity $\mu_2$ (middle panel) and about the advice accuracy $s(\mu_2)$. Black dots indicate inputs (1: helpful advice, 0: misleading advice) and colored dots simulated responses (1: going with the advice, 0: going against the advice).

### 2 Reproducibility

To ensure reproducibility of our results, we report the results excluding one participant who did not consent to make their data available for reuse and was excluded from the public repository. Our entire analysis pipeline with instructions can be found here: `https://github.com/Murdugan/compi_ioio_phase`. The data can be downloaded here: (will follow upon acceptance).

The models were implemented in Matlab (version: 2017a; `https://mathworks.com`) using the HGF toolbox (version: 3.0), which is made available as open-source code as part of the TAPAS software collection (`https://github.com/translationalneuromodeling/tapas/releases/tag/v3.0.0`; Frässle et al. (2021)) and the VBA toolbox (Daunizeau et al., 2014) (`https://mbb-team.github.io/VBA-toolbox/`; Daunizeau et al. (2014)). Statistical analyses were run in R (version: 4.04; `https://www.r-project.org/`) using R-Studio (version: 1.4.1106; `https://www.rstudio.com/`).

#### 2.1 Behavioural results without one participant

Excluding the participant who did not consent to reusing their data, we still identified a significant group-by-task-phase interaction on the frequency of advice-taking ($F = 4.857$, $p = 0.012$). To unpack this effect we repeated the analysis with three two-group models. We found significant group-by-task-phase interactions when comparing HC vs FEP ($F = 7.128$, $p_{uncorr} = 0.012$, $p = 0.035$ Bonferroni-corrected for the number of comparisons, i.e. $n = 3$) and HC vs CHR-P ($F = 7.745$, $p_{uncorr} = 0.009$, $p = 0.026$), but not when comparing CHR-P vs FEP ($F = 0.001$, $p_{uncorr} = 0.977$, $p = 1.000$), suggesting that both CHR-P and FEP showed reduced flexibility to take environmental volatility into account as the difference between stable and volatile phase was reduced compared to HC. None of the covariates significantly impacted advice taking.

The group-by-task-phase interaction remained significant after including antipsychotic and antidepressant dose as covariates ($F = 4.296$, $p = 0.019$). Neither the effect of antipsychotic dose ($F = 0.240$, $p = 0.627$) or antidepressant dose ($F = 0.096$, $p = 0.759$) were significant. Unpacking this model again revealed significant group-by-task-phase interactions when comparing HC vs FEP ($F = 7.128$, $p_{uncorr} = 0.012$, $p = 0.035$), but not when comparing CHR-P vs FEP ($F = 0.322$, $p_{uncorr} = 0.574$, $p = 1.00$). The group-by-task-phase interaction effect for HC vs CHR-P did not survive Bonferroni correction ($F = 5.154$, $p_{uncorr} = 0.030$, $p = 0.089$).

#### 2.2 Modelling results without one participant

##### 2.2.1 Bayesian model selection without one participant

We also repeated the Bayesian model selection including participants from all groups first. The results were again inconclusive ($\phi$ = 59.92%, $f$ = 51.22% in favour of Hypothesis II) possibly suggesting that different groups were best explained by different models (i.e., different computational mechanisms). To assess this possibility, we repeated the model selection for each group separately. In HC, the winning model was the standard 3-level HGF (Hypothesis I; $\phi$ = 96.63%, $f$ = 95.93%). Conversely, in FEP the mean-reverting HGF that included a drift at the third level was selected (Hypothesis II; $\phi$ = 99.89%, $f$ = 95.69%). For CHR-P, we observed a more heterogeneous results: While the mean-reverting model was favoured (Hypothesis II; $\phi$ = 84.50%, $f$ = 60.24%), there was also evidence for the standard HGF, albeit to a much lesser extent (Hypothesis I; $\phi$ = 14.41%, $f$ = 37.19%). Further inspection of the model attributions for all individual participants revealed an interesting pattern. All HC were attributed to the standard HGF with over 97% probability, whereas FEP were attributed to the mean-reverting model with over 99%. Interestingly, model attributions for CHR-P were more heterogeneous ranging from 0 to 100% probability, suggesting that some individuals were better explained by the standard HGF, but others by the mean-reverting model.

##### 2.2.2 Parameter group effects without one participant

In the reduced sample, the drift equilibrium point $m_3$ significantly differed across the groups ($\eta^2 = 0.130, p_{uncorr} = 0.030$). Post hoc tests revealed that $m_3$ was increased in FEP compared to HC suggesting that FEP perceived the intentions of the adviser as increasingly more volatile over time ($\eta^2 = 0.191, p = 0.029$, Bonferroni-corrected for the number of comparisons across groups, i.e., $n = 3$). We also performed an exploratory analysis including all other free model parameters. This analysis revealed an additional effect on coupling strength $\kappa_2$ ($\eta^2 = 0.157, p_{uncorr} = 0.014$), which was driven by reduced coupling strength between the second and third level of the perceptual hierarchy in FEP compared to HC ($\eta^2 = 0.245, p = 0.010$, Bonferroni-corrected for the number of comparisons across groups, i.e., $n = 3$). However, neither the effect on $m_3$ nor $\kappa_2$ survived Bonferroni correction for the number of parameters, i.e. $n = 7$ ($p = 0.207$ and $p = 0.100$, respectively), possibly due to a lack of power.

##### 2.2.3 Symptom-parameter correlations without one participant

Repeating the correlations on the reduced sample yielded a positive trend correlation between $m_3$ and PANSS positive symptoms ($\tau = 0.175$, $p_{uncorr} =$

0.077, $p = 0.460$ Bonferroni-adjusted for 2 (#parameters) x 3 (#PANSS subscales) = 6 comparisons). As in the main manuscript, there were negative correlations between $\kappa_2$ and PANSS negative and general symptoms ($\tau = -0.265$, $p_{uncorr} = 0.008$, $p = 0.052$ and $\tau = -0.24$, $p_{uncorr} = 0.013$, $p = 0.077$ respectively), which did not survive Bonferroni correction.

Similarly, there was only a trend correlation when excluding this subject between $m_3$ and the PCL frequency subscale ($\tau = 0.170$, $p_{uncorr} = 0.079$, $p = 0.475$ Bonferroni-adjusted for 2 (#parameters) x 3 (#PCL subscales) = 6 comparisons). Note, that the participant that did not consent to reuse of their data scored high on positive symptoms which likely contributes to the changes in the correlations compared to the full sample. However, as we pointed out in the main manuscript these correlation results should be taken as preliminary due to the small sample size of this study.
